# APOE interacts with tau PET to influence memory independently of amyloid PET

**DOI:** 10.1101/19008318

**Authors:** Alexandra J. Weigand, Kelsey R. Thomas, Katherine J. Bangen, Graham M.L. Eglit, Lisa Delano-Wood, Paul E. Gilbert, Adam M. Brickman, Mark W. Bondi, for the Alzheimer’s Disease Neuroimaging Initiative

**Affiliations:** San Diego State University/University of California, San Diego Joint Doctoral Program; Veterans Affairs San Diego Healthcare System; Department of Psychiatry, University of California, San Diego; Department of Psychology, San Diego State University; Taub Institute for Research on Alzheimer’s Disease and the Aging Brain, Department of Neurology, College of Physicians and Surgeons, Columbia University

## Abstract

**Objective:** Apolipoprotein E (APOE) interacts with AD pathology to promote disease progression. Studies of APOE risk primarily focus on amyloid, however, and little research has assessed its interaction with tau pathology independent of amyloid. The current study investigated the moderating effect of APOE genotype on independent associations of amyloid and tau PET with cognition.

**Methods:** Participants included 297 older adults without dementia from the Alzheimer’s Disease Neuroimaging Initiative. Regression equations modeled associations between cognitive domains and (1) cortical Aβ PET levels adjusting for tau PET and (2) medial temporal lobe (MTL) tau PET levels adjusting for Aβ PET, including interactions with APOE ε4 carrier status.

**Results:** Adjusting for tau PET, Aβ was not associated with cognition and did not interact with ε4 status. In contrast, adjusting for Aβ PET, MTL tau PET was significantly associated with all cognitive domains. Further, there was a moderating effect of ε4 status on MTL tau and memory with the strongest negative associations in ε4 carriers and at high levels of tau. This interaction persisted even among only Aβ negative individuals.

**Interpretation:** APOE ε4 genotype strengthens the negative association between MTL tau and memory independently of Aβ, although the converse is not observed, and this association may be particularly strong at high levels of tau. These findings suggest that APOE may interact with tau independently of Aβ and that elevated MTL tau confers negative cognitive consequences in Aβ negative ε4 carriers.

## Introduction

For the past 25 years, research on Alzheimer’s disease (AD) has been driven by an amyloid-centric model of disease pathogenesis.^1^ This model purports that AD is initiated by abnormal amyloid beta (Aβ) accumulation, later followed by tau-mediated neurofibrillary tangle formation, neurodegeneration, and downstream cognitive deficits.^2,3^ Accordingly, most research on apolipoprotein E ε4 (APOE ε4) allelic effects, as the strongest susceptibility gene for increased risk and accelerated onset of AD,^4,5^ has focused on its associations with Aβ. Research has demonstrated that APOE is involved in the oligomerization, aggregation, degradation, and clearance of Aβ,^6,7^ and that ε4 carriers have increased incidence of Aβ PET positivity as well as accelerated Aβ PET accumulation.^8,9^

Recent findings, however, also point to tau phosphorrylation as an early event in preclinical sporadic AD, possibly even preceding the involvement of Aβ.^10,11,12^ The presence of cerebral tau pathology in the absence of Aβ remains a controversial topic, with some suggesting that this is a normal age-related phenomenon when confined to the medial temporal lobe (MTL; see [13] for a review of primary age-related tauopathy, or ‘PART’) and others citing evidence that it represents an early stage on the AD continuum.^14^ A recently proposed AD diagnostic framework incorporating Aβ, tau, and neurodegeneration biomarkers (i.e., A/T/N) reinforces the primacy of Aβ over tau by labeling Aβ alone as “Alzheimer’s pathologic change” whereas tau alone is labeled “non-AD pathologic change”.^3,15^ Studies investigating tau independently of Aβ are needed to further our understanding of biomarker dynamics in preclinical AD.

Although animal studies implicate APOE in the formation of tau pathology independently of Aβ,^16,17^ there is a paucity of analogous research investigating APOE-related effects on tau measured through cerebrospinal fluid or positron emission tomography (PET) independently of one’s Aβ level. Specifically, studies investigating APOE, tau PET, and their interactive effects on cognition in Aβ negative individuals are notably lacking. Given this gap in the literature, we independently assessed associations between cognitive performance and (1) cortical Aβ PET controlling for MTL tau PET, as well as (2) MTL tau PET controlling for cortical Aβ PET, along with interactions with APOE ε4 status. For any interaction between PET and ε4 status, we also ran stratified follow-up analyses assessing these interactions separately among individuals with negativity and positivity for the other pathology (e.g., tau by APOE interaction on cognition in Aβ positive [A+] and negative [A-] individuals). We conducted these analyses with the hypothesis that presence of an APOE ε4 allele would have a deleterious moderating effect on the association between tau and cognition independent of Aβ PET level and regardless of Aβ positivity status.

## Methods

### Participants

Data used in the preparation of this article were obtained from the Alzheimer’s Disease Neuroimaging Initiative (ADNI) database (adni.loni.usc.edu). The ADNI was launched in 2003 as a public-private partnership. The primary goal of ADNI has been to test whether serial magnetic resonance imaging (MRI), positron emission tomography (PET), other biological markers, and clinical and neuropsychological assessment can be combined to measure the progression of MCI and early AD. This research was approved by the Institutional Review Boards of all participating sites, and written informed consent was obtained for all study participants. Participants without dementia (n = 212 with normal cognition; n = 89 with mild cognitive impairment [MCI]) from the ADNI were selected based on the availability of tau and Aβ PET data acquired within 12 months of each other, as well as APOE genotype data (n = 297).

### PET processing

Processing methods for ADNI Aβ PET (^18^F-AV-45, fluorbetapir) and tau PET (^18^F-AV-1451, flortaucipir) have been previously described elsewhere (http://adni.loni.usc.edu/methods/pet-analysis-method/pet-analysis/). Based on extant processing recommendations, regional standardized uptake values (SUVs) were intensity normalized using the whole cerebellum^18,19^ (Aβ PET) or inferior cerebellar gray^20,21^ (tau PET) to create SUV ratios (SUVRs). Tau PET data were partial volume corrected using the geometric transfer method.^22^ For Aβ PET, a cortical summary measure was created by averaging across FreeSurfer-derived frontal, cingulate, lateral parietal, and lateral temporal regions of interest to capture early vulnerable regions for Aβ deposition.^18^ For tau PET, a Braak stage I/II composite region was created by averaging across FreeSurfer-derived hippocampal and entorhinal regions of interest to recapitulate tau progression in early Braak stages.^20,21^

Although previous work has determined positivity thresholds via differing methods,^23-25^ positivity thresholds in this study were derived using conditional inference decision tree regression with the ctree() function from the party package in R (https://cran.r-project.org/) in order to: (1) remain consistent with prior derivations of tau PET thresholds in ADNI;^21,26^ (2) maintain comparable methods in the derivation of optimal thresholds for both Aβ and tau; and (3) independently derive thresholds for Aβ and tau, rather than determining thresholds for one based on discrimination of the other biomarker.^23,25^ Thresholds were determined using binary classification of individual SUVRs based on global cognitive function (i.e., Mini-Mental State Examination scores) and were taken at the lowest-level bifurcation in the tree, unless otherwise noted. A larger sample of individuals (n = 523) spanning all diagnostic categories (i.e., cognitively normal, MCI, dementia) with tau PET data and a subset with Aβ PET data were included for threshold derivation. A threshold of SUVR > 1.14 was determined for cortical Aβ positivity (A+/A-); although this threshold came from a higher-level bifurcation, it was selected in order to maintain consistency with other commonly used Aβ thresholds (e.g., >1.11).^18,27^ For tau PET, thresholds were first determined for higher Braak composite stages (i.e., >1.96 for stage V/VI and >1.51 for stage III/IV), with individuals surpassing the positivity threshold for higher stages iteratively removed during derivation of lower-stage thresholds, as described elsewhere.^26^ Ultimately, a threshold of SUVR > 1.18 was determined for tau Braak I/II (i.e., MTL) positivity (T+/T-), largely consistent with a previously derived threshold using similar methods (>1.13).^21^

### Cognitive testing

Participants underwent comprehensive neuropsychological testing including the measures from the following domains: attention/executive function (Trail Making Test Parts A and B, time to completion); language (confrontation naming [i.e., Boston Naming Test or Multilingual Naming Test]and animal fluency); and memory recall (Logical Memory Story A Immediate and Delayed Recall). All raw scores were converted to z-scores based on predicted values from regression equations adjusting for age, sex, and education derived within a robust normal control group (i.e., cognitively normal throughout their duration in ADNI) based on the entire ADNI sample. Z-scores were then averaged within domains to create attention/executive, language and memory composite scores. MCI was diagnosed using actuarial neuropsychological criteria.^28,29^ Participants were diagnosed with MCI if they (1) had two impaired scores in one cognitive domain or (2) had one impaired score across all three cognitive domains.

### APOE genotyping

All participants had APOE ε4 genotyping data available. ε4 carriers and non-carriers were determined based on presence of at least one ε4 allele. Of the 297 participants overall, 99 (33%) were categorized as ε4 carriers (heterozygotes n = 82; homozygotes n = 17) and 198 (67%) as non-carriers.

### Statistical analyses

Cognitive domain z-scores were shifted to a positive scale and Box-Cox transformed to improve normality. Tau PET SUVRs were also Box-Cox transformed to improve normality. All figures depict untransformed values to facilitate interpretation. Chi-squared tests and t-tests assessed for differences in demographic variables, PET values, and cognitive composite scores between ε4 carriers and non-carriers. Regression equations predicted demographically-adjusted and Box-Cox transformed cognitive z-scores as a function of cortical Aβ PET SUVRs while controlling for MTL tau PET SUVRs (model 1) and MTL tau PET SUVRs while controlling for cortical Aβ PET SUVRs (model 2). Both models included linear and quadratic SUVR-of-interest main effects controlling for APOE ε4 status and, in separate models, interactions with APOE ε4 status. For any significant linear or quadratic interactions, follow-up analyses examined these interactions stratified by PET positivity status (e.g., tau PET and ε4 interactions for separately for A- and A+ individuals). All analyses and figures were generated in R version 3.5.0 (https://cran.r-project.org/).

## Results

### Sample characteristics

Group differences between ε4 carriers and non-carriers are presented in Table 1. Participants’ sex distribution and years of education did not differ between carriers and non-carriers, but ε4 carriers were younger. APOE ε4 carriers had a higher average cortical Aβ SUVR and a higher proportion of Aβ positive individuals relative to non-carriers. Further, ε4 carriers had a higher average MTL tau SUVR relative to non-carriers, although the proportion of tau positive individuals was similar between groups. APOE ε4 carriers and non-carriers did not differ in any cognitive domain.

**Table 1.** APOE ε4-carrier and non-carrier group differences in demographic and PET variables.

|  | **ε4 carriers**<br>**(n=99)** | **Non-carriers**<br>**(n=198)** | **Test statistic** | **p-value** |
| --- | --- | --- | --- | --- |
| **Age (Mean[SE])** | 74.28(.72) | 76.50(.52) | t = 2.50 | p = .01 |
| **Sex (% female)** | 53.5 % female | 46.5% female | $\chi^2 = 1.05$ | p = .30 |
| **Education (Mean[SE])** | 16.51(.27) | 16.83(.17) | t = 1.05 | p = .29 |
| **Aβ status (% A+)** | 50.5% A+ | 23.7% A+ | $\chi^2 = 20.30$ | p < .001 |
| **Tau status (% T+)** | 72.7% T+ | 71.7% T+ | $\chi^2 = .002$ | p = .96 |
| **Cortical amyloid**<br>**SUVR (Mean[SE])** | 1.21(.02) | 1.09(.01) | t = 5.07 | p < .001 |
| **MTL tau SUVR**<br>**(Mean[SE])** | 1.39(.03) | 1.32(.02) | t = 2.13 | p = .03 |
| **Attention/executive**<br>**(Mean[SE])** | -.45(.13) | -.25(.08) | t = 1.37 | p = .17 |
| **Language (Mean[SE])** | -.46(.14) | -.38(.09) | t = .49 | p = .62 |
| **Memory (Mean[SE])** | -.65(.14) | -.43(.08) | t = 1.41 | p = .16 |

### APOE by Aβ interaction on cognition

In models adjusting for tau PET SUVR and ε4 status, there was no linear or quadratic main effect of Aβ on cognitive performance in any domain (all *t*s < |1.44|, *p*s > .15). Further, no moderating effect of ε4 status was observed for the linear and quadratic associations between cortical Aβ SUVRs and any cognitive domain (all *t*s < |1.24|, *p*s > .22) while controlling for MTL tau PET SUVR.

### APOE by tau interaction on cognition

In models adjusting for cortical Aβ PET SUVR and ε4 status, there was a linear main effect of MTL tau for attention/executive performance (β = -.21, *t* = −3.46, *p* < .001) such that higher levels of tau were associated with poorer attention/executive performance. There was also a quadratic main effect of MTL tau for language (β = -.25, *t* = −3.46, *p* < .001) and memory (β = −1.09, *t* = −2.45, *p* < .001) such that higher levels of tau were associated with worse performance and this negative association was disproportionately stronger at higher levels of tau. Further, a moderating effect of ε4 status was observed for the association between quadratic MTL tau SUVRs and memory performance (β = .73, *t* = 3.54, *p* < .001) such that ε4 carriers exhibited a disproportionately stronger negative association between tau and memory at higher levels of tau (see Figure 1). Notably, the moderating effect of ε4 status on quadratic MTL tau-memory associations was upheld even among individuals who had not yet reached Braak stage III/IV (β = .49, *t* = 2.91, *p* = .004). There were no moderating effects of ε4 status for the associations between linear or quadratic MTL tau and language or attention/executive performance (all *t*s < |1.59|, *p*s > .11).

**Figure 1.**
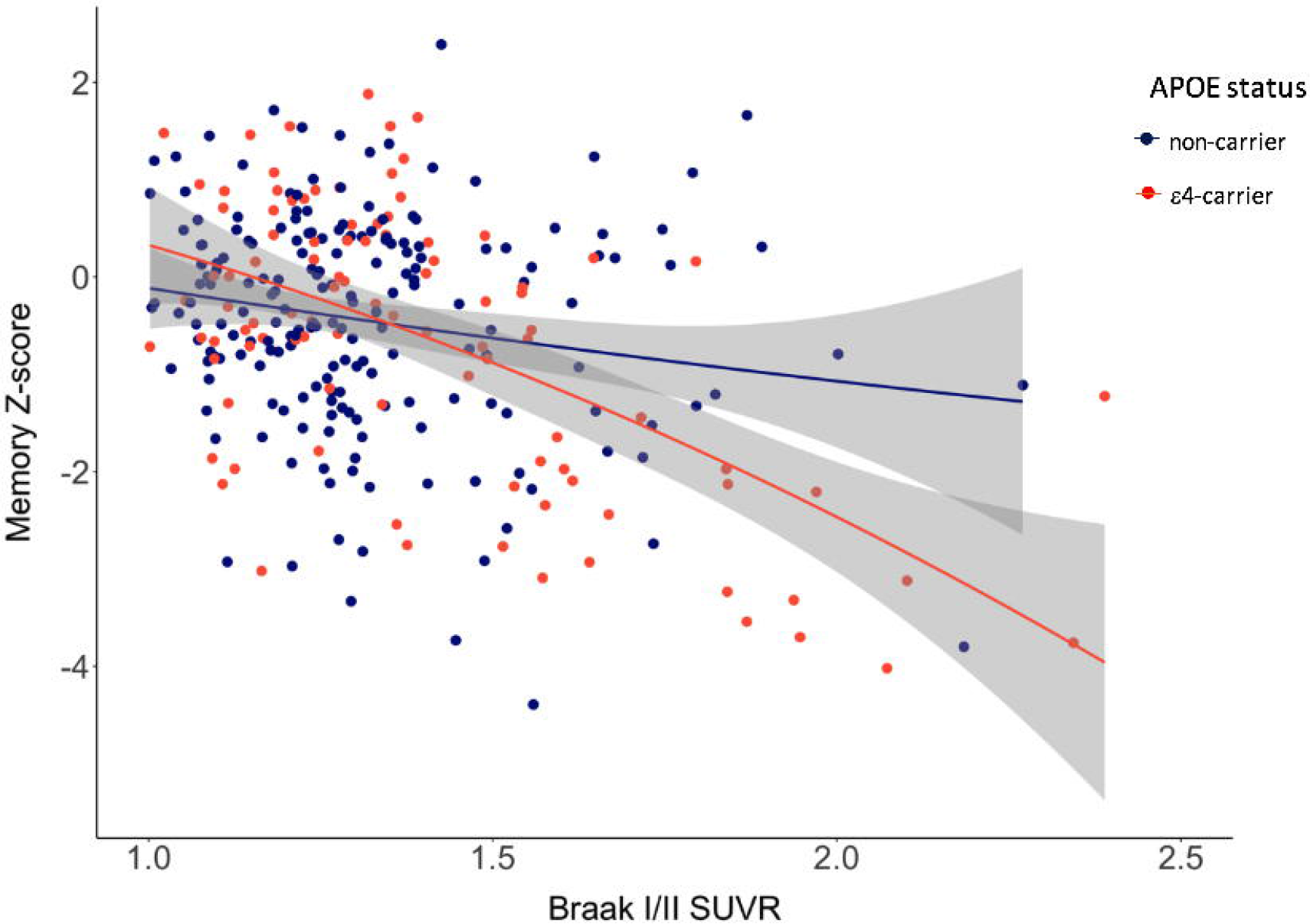
Quadratic moderating effect of ε4-carrier status on the association between medial temporal lobe tau and memory performance.

Follow-up stratified analyses were conducted for the interaction between APOE ε4 status and MTL tau level on memory performance to examine the effects separately for Aβ negative (A-) and positive (A+) individuals. Among A+ individuals, a moderating effect of ε4 status on linear MTL tau and memory performance was observed (β = 1.02, *t* = 2.17, *p* = .03) such that higher levels of tau were more strongly associated with poor memory performance among ε4 carriers (see Figure 2A). However, unlike in the model with the full sample, no interaction was observed for the quadratic effect of MTL tau in this A+ group (β = .07, *t* = .79, *p* = .43). In contrast, among A-individuals, a moderating effect of ε4 status on quadratic MTL tau and memory performance was observed (β = .20, *t* = 2.12, *p* = .03) such that a negative association between tau and memory emerged only for higher levels of tau among ε4 carriers (see Figure 2B).

**Figure 2.**
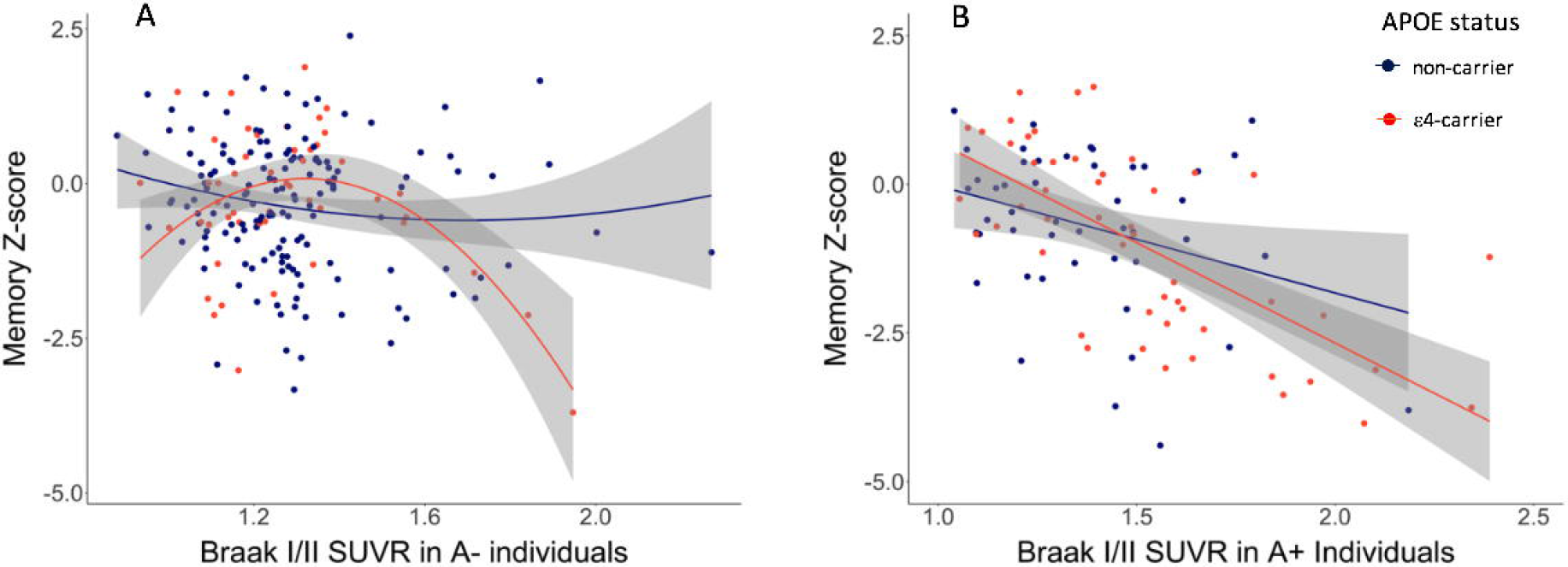
A: Quadratic moderating effect of ε4-carrier status on the association between medial temporal lobe tau and memory performance among amyloid beta negative individuals. B: Linear moderating effect of ε4-carrier status on the association between medial temporal lobe tau and memory performance among amyloid beta positive individuals.

## Discussion

Moderating effects of ε4 status on the association between AD PET biomarker levels and cognition were observed only for associations between tau PET and memory, such that higher levels of tau were associated with poorer memory performance only among ε4 carriers, and this effect was disproportionately strong at higher levels of tau. Notably, this interaction was observed independently of Aβ PET level as measured on a continuous scale and among individuals who were Aβ negative via a threshold method. Further, independently of Aβ PET level and ε4 status, tau PET was negatively associated with both language and attention/executive performance. In contrast, when adjusting for tau PET level, no main effects of Aβ PET or moderating effects of ε4 status on cognition were observed. These findings suggest that tau interacts with APOE ε4 independently of Aβ to exert negative influences on cognition, warranting a primary role for tau within the preclinical AD framework.^3,30^

As a consequence of the amyloid cascade model, the majority of research on AD over the past few decades has focused on the role of Aβ in AD^1^ or, at the very least, investigated other pathologic markers in the context of Aβ positivity—but less so among those with Aβ negativity. Although a more recent model of AD pathogenesis known as the A/T/N framework^31^ has expanded its purview and purports agnosticism to the temporal sequencing of mechanisms underlying AD pathogenesis, it still relegates tau in the absence of Aβ to an alternative categorization of “non-AD pathologic change.” In contrast to these amyloid-centric models, there is accumulating evidence in support of a continuum hypothesis in which tau pathology accumulates in the brainstem and propagates to transentorhinal cortex independently of and prior to Aβ.^11,32,33^ Only cases with tau positivity in the absence of Aβ and clinical dementia would warrant “non-AD pathologic change,” but individuals in this pathologic category without dementia are considered on the Alzheimer’s continuum.^14^ Additionally, recent research consistently shows strong associations between tau PET and cognition that are not evident with Aβ PET.^34-36^ Our findings of a moderating effect of APOE ε4 status on tau and memory associations independently of Aβ, which persists among Aβ negative individuals, build upon these prior studies to provide further support for the notion that tau has a role in AD nosology independent from Aβ.

The mechanisms by which APOE ε4 exerts its deleterious effects are incompletely understood, although both Aβ-dependent and -independent avenues have been explored. In addition to its role in the aggregation and clearance of Aβ, APOE has been implicated in neuroinflammatory processes, cerebrovascular alterations, and synaptic plasticity,^37^ but its direct and indirect effects on tau pathology remain unclear. Evidence suggests that misfolding and aggregation of tau may be directly influenced by APOE-relevant processes.^17^ Further, a mouse model of tauopathy expressing different APOE genotypes demonstrated that ε4 knock-in mice had, in addition to higher tau levels, higher microglial reactivity and tumor necrosis factor alpha (TNF-α) secretion, indicating an ε4-induced neuroinflammation independent of Aβ.^16^ Thus, our findings may reflect possible APOE-mediated inflammatory processes exacerbating tau-mediated neurodegeneration, resulting in the observed associations with cognition.

Given findings that APOE regulates clearance of Aβ pathology,^38,39^ it is reasonable to speculate that it may be involved in the clearance of tau pathology as well. Although pathologic tau aggregates intraneuronally to form large fibrillar structures known as tangles, smaller soluble oligomers exist both intra- and extra-cellularly. These toxic oligomeric tau species form prior to tangles, spreading through neighboring cells in a transsynaptic process to seed the aggregation of intraneuronal tangles.^40^ Soluble oligomeric tau species may exacerbate inflammatory processes41 as well as inhibit post-synaptic proteins, reduce gliotransmitter release, and impair long-term potentiation and memory.^42,43^ Reduced clearance of these tau oligomers in ε4 carriers may potentiate these negative effects, resulting in the observed strengthened associations between tau and memory performance among ε4 carriers.

When stratifying the tau-memory associations based on Aβ positivity, differential patterns of ε4 moderation were observed. Among Aβ negative individuals, ε4 carriers exhibited a quadratic effect such that a negative association between tau and memory emerged only at higher levels of tau. In contrast, among Aβ positive individuals, a linear interaction was observed such that the negative association between all levels of tau and memory was stronger in ε4 carriers. It is conceivable that, in the context of high Aβ, tau is associated with poorer memory even at relatively lower tau levels because the combined effects of Aβ and ε4 status on neuroinflammatory processes lower the threshold such that less tau is needed to initiate neurodegeneration and cognitive dysfunction. Instead, when Aβ levels fall below the positivity threshold, a higher amount of tau pathology in combination with ε4-associated processes may be needed to exert a negative effect on memory, explaining the observed quadratic effect among Aβ negative individuals. Notably, the strongest negative effects of tau on memory emerged for both groups soon after the SUVR had surpassed the Braak I/II positivity threshold, indicating a potentially important threshold effect for tau regardless of Aβ status. Further, given the quadratic nature of the interaction such that effects on memory were most prominent at higher levels of MTL tau, analyses were rerun among only individuals negative for Braak stage III/IV; the ε4 interaction on quadratic MTL tau-memory associations was retained, suggesting that the strengthened effect for high MTL tau is not driven by individuals who have reached Braak stage III/IV.

The primary strength of this study was the investigation of the moderating effects of APOE ε4 on both Aβ and tau PET independently, resulting in the novel finding that the quadratic association between tau PET and memory was retained in ε4 carriers even among Aβ negative individuals. Further, assessing multiple cognitive domains broadened the scope of investigation beyond memory to demonstrate robust main effects of MTL tau on language and attention/executive functions independently of Aβ level. However, generalizability is limited given that this study was conducted in a racially/ethnically homogenous, clinic-based sample with few medical comorbidities. Replication of these findings in a more representative sample will better inform the range of pathologic and associated cognitive changes in AD. Additionally, extending analyses to other regions of tau deposition and other pathologies such as TDP-43 and cerebrovascular changes will provide more insight into the polypathologic nature of AD and the widespread influence of APOE genotypic variations on disease expression.

Our findings suggest that APOE may exert deleterious effects on cognition through specific interactions with tau pathology and that these effects may occur independently of and prior to the amyloidosis of AD. This relationship has important implications for models of AD pathogenesis by supporting an Aβ-independent role of APOE and tau during the preclinical period of AD that is further amplified in the presence of high Aβ.

## Data Availability

Data used in the preparation of this article were obtained from the Alzheimer’s Disease Neuroimaging Initiative (ADNI) database (adni.loni.usc.edu). The ADNI was launched in 2003 as a public-private partnership. The primary goal of ADNI has been to test whether serial magnetic resonance imaging (MRI), positron emission tomography (PET), other biological markers, and clinical and neuropsychological assessment can be combined to measure the progression of MCI and early AD. This research was approved by the Institutional Review Boards of all participating sites, and written informed consent was obtained for all study participants.

http://adni.loni.usc.edu/

## Acknowledgments

This work was supported by NSF fellowship DGE-1650112 (A.J.W.) and NIH grants R01 AG049810 and R01 AG054049 (M.W.B.). Data collection and sharing for this project was funded by the Alzheimer’s Disease Neuroimaging Initiative (ADNI) (National Institutes of Health Grant U01 AG024904) and DOD ADNI (Department of Defense award number W81XWH-12-2-0012). ADNI is funded by the National Institute on Aging, the National Institute of Biomedical Imaging and Bioengineering, and through generous contributions from the following: AbbVie, Alzheimer’s Association; Alzheimer’s Drug Discovery Foundation; Araclon Biotech; BioClinica, Inc.; Biogen; Bristol-Myers Squibb Company; CereSpir, Inc.; Cogstate; Eisai Inc.; Elan Pharmaceuticals, Inc.; Eli Lilly and Company; EuroImmun; F. Hoffmann-La Roche Ltd and its affiliated company Genentech, Inc.; Fujirebio; GE Healthcare; IXICO Ltd.; Janssen Alzheimer Immunotherapy Research & Development, LLC.; Johnson & Johnson Pharmaceutical Research & Development LLC.; Lumosity; Lundbeck; Merck & Co., Inc.; Meso Scale Diagnostics, LLC.; NeuroRx Research; Neurotrack Technologies; Novartis Pharmaceuticals Corporation; Pfizer Inc.; Piramal Imaging; Servier; Takeda Pharmaceutical Company; and Transition Therapeutics. The Canadian Institutes of Health Research is providing funds to support ADNI clinical sites in Canada. Private sector contributions are facilitated by the Foundation for the National Institutes of Health (www.fnih.org). The grantee organization is the Northern California Institute for Research and Education, and the study is coordinated by the Alzheimer’s Therapeutic Research Institute at the University of Southern California. ADNI data are disseminated by the Laboratory for Neuro Imaging at the University of Southern California.

## Author Contributions

AJW: conception and design of the study, acquisition and analysis of data, drafting manuscript and/or figures

KRT: conception and design of the study, revising the manuscript for intellectual content

KJB: conception and design of the study, revising the manuscript for intellectual content

GMLE: conception and design of the study, revising the manuscript for intellectual content

LDW: conception and design of the study, revising the manuscript for intellectual content

PEG: conception and design of the study, revising the manuscript for intellectual content

AMB: conception and design of the study, revising the manuscript for intellectual content

MWB: conception and design of the study, acquisition and analysis of data, revising the manuscript for intellectual content

